# Feasibility of a Community Health Worker-delivered Tobacco Cessation Intervention During Pregnancy: Protocol for the Smoke-Free HOPE Pilot Randomized Controlled Trial

**DOI:** 10.1101/2025.10.26.25338756

**Authors:** Anne Berit Petersen, Berenice Rosas, Juan Carlos Belliard, Janice Tsoh, Bryan Oshiro, Ruofan Yao, Pramil N. Singh

## Abstract

**Background:** In the US, maternal tobacco use is more prevalent in pregnant individuals reporting less than a high school education, low income, rural residence, no private insurance coverage, a mental health diagnosis, and/or race/ethnicity as being American Indian/Alaska Native, Black/African American, or White. Prior evaluation of a large state-funded multi-component maternal smoking cessation program in San Bernardino County demonstrated strong short-term abstinence but substantial dropout and relapse, highlighting the need for alternative delivery models.

**Objective:** To address these challenges, the **Smoke-Free HO**usehold and **P**ar**E**nt (*Smoke-Free HOPE*) protocol was developed to deliver tobacco cessation support through community health worker (CHW) visits. This study protocol describes a pilot randomized controlled trial designed to evaluate the feasibility and acceptability of *Smoke-Free HOPE*, a CHW-delivered tobacco cessation intervention tailored for pregnant individuals.

**Methods:** This protocol uses a sequential mixed-methods design under a Capability, Opportunity, Motivation-Behavior (COM-B) framework. The study includes (a) a stakeholder analysis for intervention design, and (b) a two-arm pilot feasibility randomized controlled trial enrolling 30 pregnant individuals who want to quit habitual use of tobacco products. The stakeholder analysis will be conducted through a Community Advisory Board. Participants will be randomized to one of two CHW-led models that deliver home or telehealth visits: (a) CHW-Brief Advice (CHW-BA) delivers a maternal smoking cessation curriculum (SCRIPT™) adapted for CHWs and cessation of all tobacco products, and (b) CHW-Patient Navigation (CHW-PN) delivers patient navigation and ongoing support for enrollment and engagement using a CHW-curated list of community-based cessation programs. Primary outcomes will assess feasibility (recruitment, retention, and engagement) and acceptability (participant satisfaction, perceived support, and motivation to quit), alongside qualitative evaluation of participant, CHW, and provider experiences; workflow integration; and behavioral and contextual barriers and facilitators influencing implementation and engagement.

**Results:** The study was funded by the Tobacco-Related Disease Research Program (TRDRP), administered by the University of California Office of the President. Participant recruitment occurred over an 18-month period between 2024 and 2026 and concluded in May 2026. Follow-up activities are nearing completion, while data management and preliminary analyses are underway. Study findings are expected to be submitted for publication in 2026.

**Conclusions:** This protocol evaluates two CHW-delivered models for maternal tobacco cessation tailored to pregnancy. Intervention components were selected to address individual, social, and structural barriers to maternal tobacco cessation. Findings will inform refinement of intervention components, implementation strategies, and outcome selection for a future definitive trial.

**Trial registration:** ClinicalTrials.gov NCT06438549

**Protocol version:** Version 2.0

## 1 Introduction

### 1.1 Background

The habitual use of tobacco products remains the most prevalent form of substance use during pregnancy [1]. The 2014 U.S. Surgeon General’s Report concluded that habitual use of tobacco products harms both maternal and fetal health and contributes to adverse birth outcomes [2]. To date, the most consistent risk factor for maternal tobacco use in the US is educational attainment of less than high school [3–5]. Other risk factors include household income, rurality, Medicaid member, depression, and those identifying as American Indian/Alaska Native, Black/African American, or White [6–8].

San Bernardino County, CA, has long reported one of the highest infant mortality rates in the state, particularly in rural areas of the county and among non-Hispanic Black infants [9, 10]. From 2012 to 2019, our group at Loma Linda University Health (LLUH) ran the state-funded (First5CA.gov) Comprehensive Tobacco Treatment Program (CTTP), the largest prenatal smoking cessation program in San Bernardino County, which health educators primarily implemented in a classroom setting [11]. Our 2022 program evaluation of CTTP participant outcomes (n=1402) indicated that, despite achieving a high rate of prolonged abstinence (40.1 %) during the 8-week program, CTTP still reported a high dropout rate (44% overall) [11]. Another noteworthy program weakness was that program participants from racial/ethnic groups with the highest national rates of tobacco use during pregnancy (American Indian/Alaskan Native, Black/African American, White) also had the highest rate of relapse during CTTP [12]. Lastly, CTTP follow-up predated the major increase in the availability of e-cigarettes/vape products. It thus cannot provide insight into how these products affect outcomes in tobacco cessation programs [12].

Based on this evaluation, the protocol for the **Smoke-Free HO**usehold and **P**ar**E**nt (*Smoke-Free HOPE*) pilot feasibility trial was developed to address three unmet needs identified in prior maternal tobacco cessation programming [11, 12]. First, the protocol addresses low retention rates by moving from classroom or clinical settings to telehealth and home visits with CHWs. Second, the protocol considers the complex tobacco exposome that includes non-combustible products (e-cigarette and vaping products, IQOS, oral nicotine pouches). Third, our model accounts for the evidence indicating that tobacco use during pregnancy tends to occur in a socioecological framework of other risk factors such as poor educational attainment, poverty (i.e., food insecurity, housing instability), and use of other substances (i.e., cannabis, opioids) [4, 5, 13, 14]. To address these gaps, the *Smoke-Free HOPE* protocol will develop and test the first CHW-delivered tobacco cessation intervention tailored for pregnant individuals in the US.

The *Smoke-Free HOPE* protocol will include an intervention development phase (i.e., using evidence-based components with known efficacy from CTTP and obtaining input from a Community Advisory Board (CAB)) and an intervention testing phase in which we conduct a pilot feasibility randomized controlled trial. The feasibility objectives for the pilot feasibility RCT include: (a) Examine the feasibility of CHW-delivered behavioral intervention models (CHW-delivered brief advice, CHW-delivered patient navigation) tailored to pregnant individuals in a two-arm RCT; (b) Examine the acceptability of CHW-delivered behavioral intervention models tailored to pregnant individuals in a two-arm RCT.

## 2 Methods

### 2.1 Overview of Study Design

*Smoke-Free HOPE* is a two-arm pilot RCT using a sequential mixed-methods design [15–17] to assess the feasibility and acceptability of two CHW-led cessation models in pregnancy: Brief Advice (CHW-BA) and Patient Navigation (CHW-PN). The study has two phases: (a) CAB development and stakeholder interviews to inform the intervention, (b) pilot RCT testing with outcomes obtained from surveys and interviews analyzed under the COM-B Framework (domains: Capability, Opportunity, Motivation [18]) to examine participant experiences and contextual barriers and facilitators.

The pilot RCT is registered at ClinicalTrials.gov (NCT06438549). Reporting follows the SPIRIT checklist and the CONSORT extension for pilot and feasibility trials [19–21]. The SPIRIT pilot randomized controlled trial protocol checklist is attached as Multimedia Appendix 1. Table 1 presents the SPIRIT diagram summarizing the enrolment, intervention delivery, and outcome assessment schedules for both the pilot RCT and the process evaluation. Pre-specified progression criteria related to feasibility outcomes will guide progression to a future definitive trial.

**Table 1.**
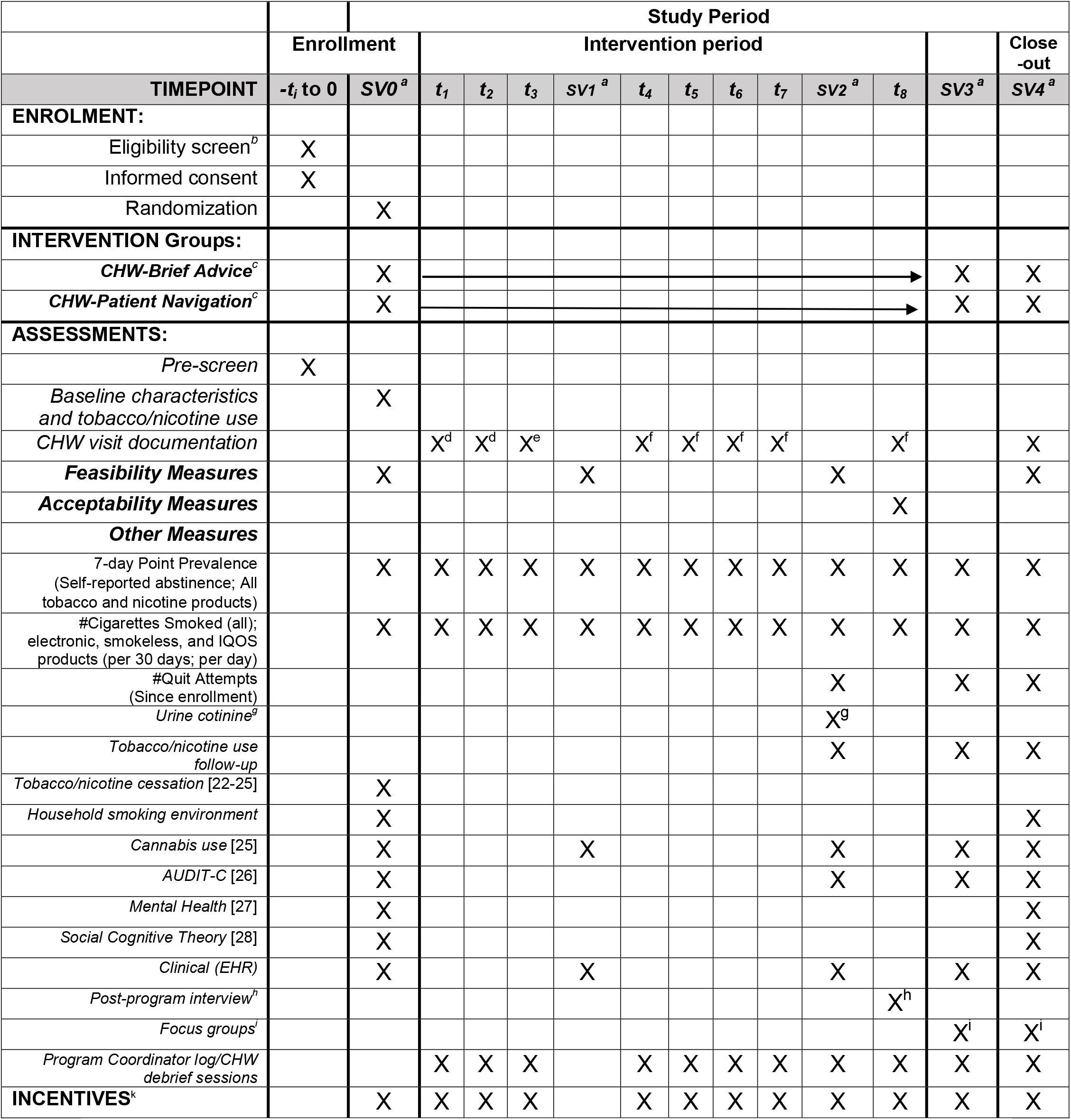
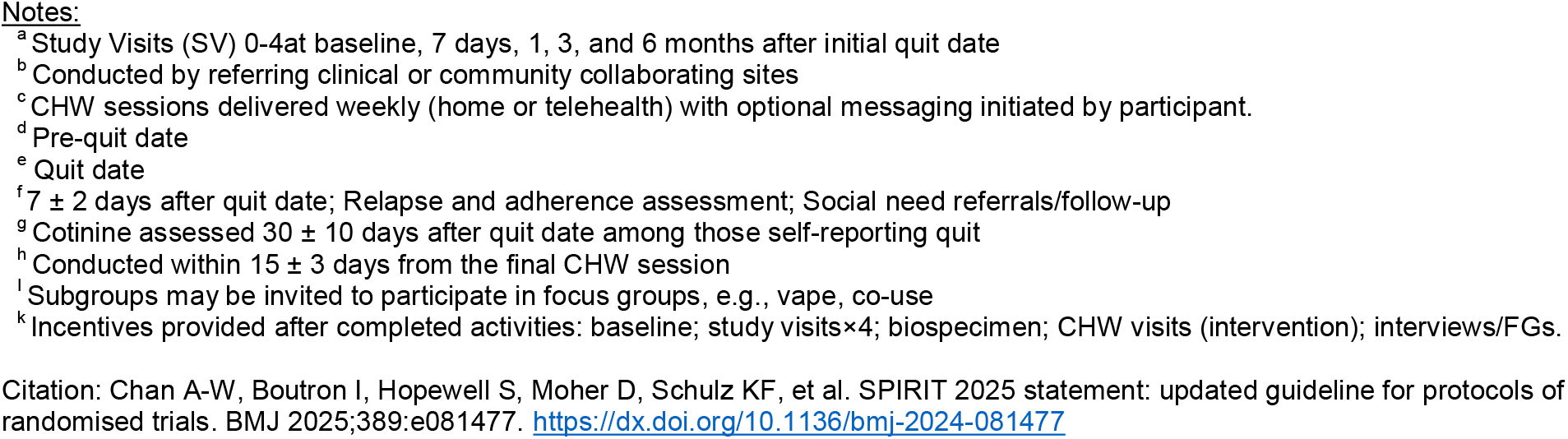
SPIRIT Schedule of enrollment, interventions, and assessments for Smoke-Free HOPE clinical trial.

### 2.2 Study Setting

The study setting includes recruitment from the Inland Empire region, which comprises 11% of the CA population [29] including: (1) the Maternal-Fetal Medicine Department at LLUH receives referrals from Federally Qualified Health Centers (FQHCs) in San Bernardino County and uses a multidisciplinary team to support referrals to social determinants of health (SDOH) programs; and (2) a referral network of prenatal care provider clinics and community-based organizations in San Bernardino and the surrounding Inland Empire region. The study team conducts data coordination with support from the LLUH Center for Data Science.

### 2.3 Participant Eligibility

The trial aims to recruit pregnant adults (≥18 years) residing in San Bernardino or Riverside Counties who are current habitual users of commercial tobacco products or who have recently quit and wish to remain abstinent. Current use includes: (a) combustible cigarette users with a lifetime history of at least 100 cigarettes and who report smoking more than three cigarettes, cigars, or blunts per day within the past seven days; (b) users of e-cigarettes/vapes containing a tobacco component on at least 25 of the past 30 days; (c) users of heated tobacco products (e.g., IQOS™) on at least 25 of the past 30 days; (d) users of smokeless tobacco on at least 25 of the past 30 days; or (e) poly-tobacco users who meet the 25-of-30-day threshold with any combination of the above products. Cessation is defined as quitting the use or poly-use of all current commercial tobacco products of choice [i.e., combustible, e-cigarettes/vaping, other smokeless tobacco products]. Sensitivity analyses will consider other cessation outcomes involving replacing combustible products with smokeless nicotine delivery products [e.g., e-cigarettes, iQOS, oral nicotine pouches].

Eligible participants must either (a) be willing to quit, (b) set a quit date within two weeks, or (c) have quit during the current pregnancy (including self-directed nicotine replacement therapy [NRT]) but be concerned about relapse. Individuals using NRT at baseline will be excluded; however, participants who initiate or continue NRT as part of usual clinical care during the study will not be discouraged from doing so.

Exclusion criteria include: (a) severe mental health conditions precluding informed consent or CHW intervention participation; (b) participation in another cessation program within the past 30 days; or (c) unwillingness to participate in audio-recorded interviews or focus groups. These criteria balance representativeness with intervention suitability and the needs of outcome assessment.

### 2.4 Recruitment

Recruitment for the *Smoke-Free HOPE* trial will use a multi-pronged strategy (self-directed referral, provider referral pathways, electronic health records, and community partnerships) designed to maximize outreach and outlined as described below.

#### 2.4.1 Self-directed referrals

These referrals primarily occur through our placement of recruitment materials (e.g., electronic and printed flyers with QR codes linking to the study website) and through links to the study website at health care sites, maternal health network sites, and in media announcements.

#### 2.4.2 Provider Referral Pathways

These on-site referrals occur during routine prenatal care at LLUH. As part of standard prenatal intake, pregnant patients are asked about tobacco product use by members of the prenatal care team, such as a nurse or social worker. Patients who report tobacco use may then be referred to *Smoke-Free HOPE*.

#### 2.4.3 Electronic Health Record

Electronic Health Record (her) informatics (MyChart, Epic Systems Corp.), will be used to identify pregnant individuals who reported tobacco or e-cigarette/vape pen use within the past three months. Eligible patients will receive tailored outreach messages through the EHR patient portal with the option to indicate interest in the study by selecting “I’m interested”.

#### 2.4.4 Community Partnerships

The referral network will expand through partnerships with community-based organizations that enable outreach at maternal health and perinatal events, presentations to local organizations and coalitions, and collaboration with regional health plans to facilitate referrals. All strategies will be refined in an iterative process in collaboration with the CAB.

### 2.5 Enrollment and Consent Procedures

The *Smoke-Free HOPE* trial will use a two-step informed consent process. During a brief telephone/video pre-screen, the Program Coordinator (PC) will obtain verbal permission to assess preliminary eligibility. Before any study activities, eligible participants will provide written informed consent with HIPAA authorization and acknowledgment of the California Experimental Research Subject’s Bill of Rights. Consent will typically be completed via secure video (or in person), with key study elements summarized. Executed copies will be provided to participants, and the consent process will be documented in accordance with institutional and regulatory requirements. Enrollment will be finalized when the site Medical Director (or clinical designee) reviews the record and signs off that all eligibility criteria are met.

### 2.6 Randomization and Blinding

A study statistician generated the allocation sequence using permuted block randomization in SAS (PROC PLAN; SAS Institute Inc.). Following enrollment, the program coordinator assigned each participant a unique study identification number and allocated the participant to the corresponding study group according to the prespecified randomization schedule. Because the intervention is behavioral, participants and CHWs cannot be blinded to group assignment; however, outcome assessors and data analysts will remain blinded to allocation to minimize bias.

### 2.7 Incentives

Participants will receive prorated Amazon e-gift cards tied to specific study activities (See Table 1). Additional compensation is offered for participation in qualitative activities (interviews and focus groups), and a final incentive is provided at the 6-month study follow-up. Participants can receive up to $300 in total compensation for all study activities.

Incentives are provided for participation in the program and each activity. Incentives are issued upon completion of each activity and are not contingent on cessation outcomes. Total compensation possible has been calibrated to align with typical U.S. behavioral trials in pregnant populations and ethical guidance to avoid undue influence [30].

### 2.8 Community Advisory Board Engagement

CAB input was a central component of the development of the *Smoke-Free HOPE* protocol. CAB engagement was grounded in community-based participatory research principles, including shared decision-making, co-learning, mutual respect, and transparency [31].

The CAB includes representatives from the San Bernardino County Department of Public Health, Community and Family Health Services; Black Infant Health of San Bernardino; First 5 San Bernardino; the Maternal Health Network of San Bernardino; and the Sankofa Birth Collective, representing public health, perinatal, early childhood, and birth equity perspectives.

Between 2022 and 2024, CAB members provided iterative input through structured meetings to refine intervention components, recruitment strategies, study procedures, and feasibility and acceptability outcomes. The team documented, synthesized, and shared feedback with CAB members to support transparency and ensure alignment with community priorities.

CAB input directly informed adaptation of the e-cigarette and dual-use cessation component, refinement of CHW training materials, and tailoring of intervention messaging to address pregnancy-specific social and structural barriers.

Overall, CAB engagement has informed intervention development during the protocol phase and is intended to support implementation refinement and dissemination planning, consistent with community-based participatory research principles. CAB members receive honoraria consistent with institutional policy and community-engaged research best practices.

### 2.9 Interventions

#### 2.9.1 Theoretical Framework for Interventions

The *Smoke-Free HOPE* intervention is grounded in Social Cognitive Theory (SCT) (48) and the NIMHD Research Framework (socioecological model) (49), which together guide the intervention’s behavioral and structural components. Guided by these frameworks, CHWs deliver home/telehealth visits using dual guidance styles: (a) directive cessation support and (b) non-directive assistance for social needs, each mapped to specific SCT and NIMHD domains [32, 33].

Directive support targets self-efficacy, behavioral capability, and outcome expectancies, while non-directive support addresses socio-cultural and environmental influences. Together, these approaches operationalize SCT’s concepts of reciprocal determinism, cessation behaviors, and the household environment (See Figure 1).

**Figure 1:**
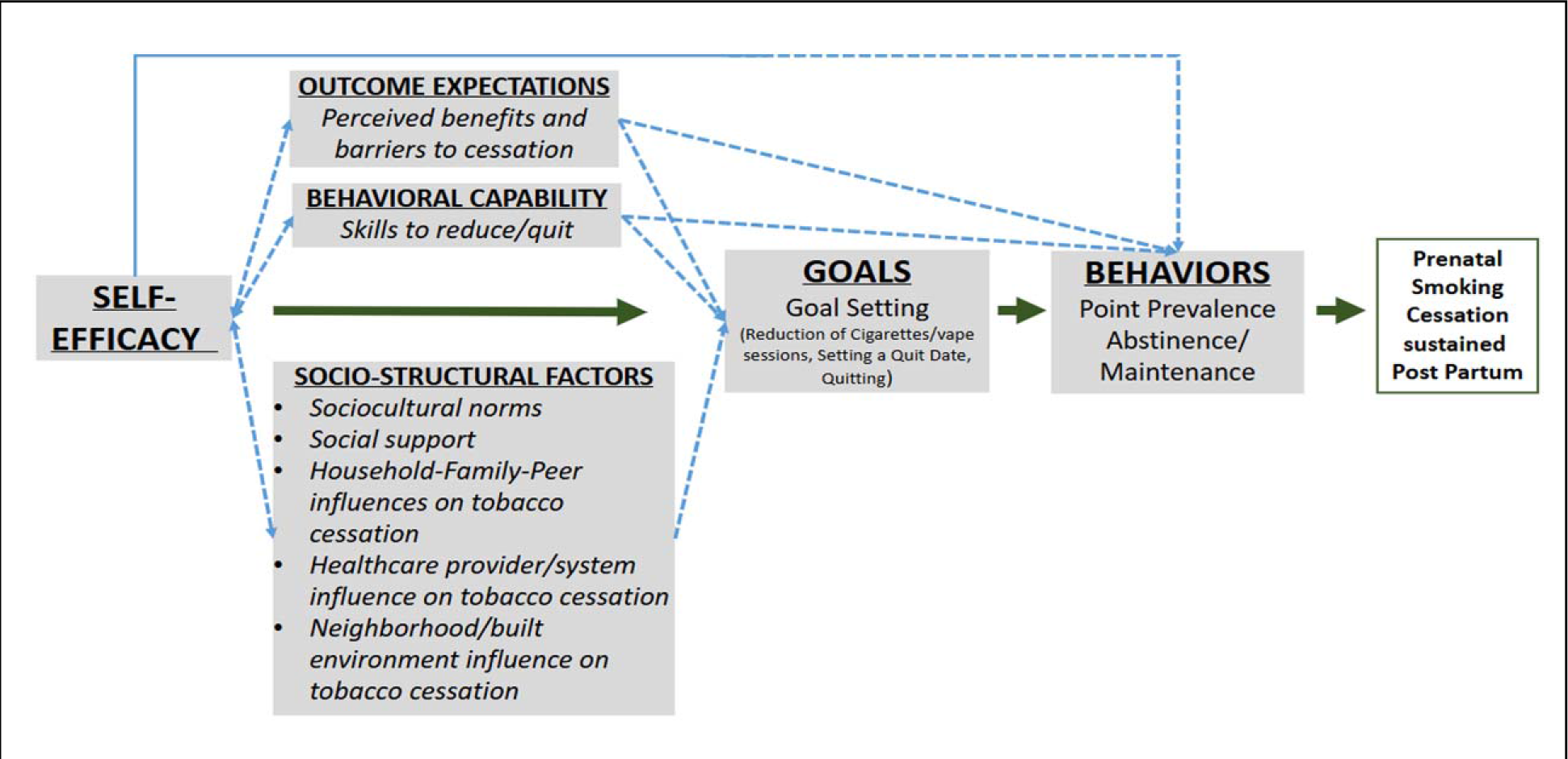
Social Cognitive Theory (SCT)-Guided CHW Support for Maternal Tobacco Cessation. CHWs partner with pregnant individuals and their families to support tobacco cessation through coordinated, interrelated mechanisms. During home or telehealth visits, CHWs (1) strengthen outcome expectancies by increasing awareness of perceived benefits and addressing personal and environmental barriers (e.g., establishing smoke-free household rules, engaging partners/household members in support); (2) build behavioral capability by facilitating access to cessation skills and practical strategies; (3) shape socio-structural factors by reinforcing clinical guidance, leveraging family/peer support, and addressing broader influences (social needs and neighborhood context); (4) facilitate goals setting by assisting with planning quit dates, and providing relapse-prevention support; and (5) enhance self-efficacy through ongoing encouragement and problem-solving. Together, these mechanisms promote sustained cessation behaviors from pregnancy through postpartum maintenance.

Within this framework, intervention delivery incorporates strategies that promote outcome expectancies (e.g., identifying benefits and barriers to cessation), build cessation-related skills and resource awareness, shape the home environment (e.g., establishing smoke-free rules and engaging household support), facilitate goal setting and quit planning, and reinforce self-efficacy through ongoing encouragement and problem-solving.

The intervention also operationalizes the NIMHD Research Framework by targeting individual and interpersonal (personal and family functioning), environment (personal and household), sociocultural (family networks), and healthcare system levels. The delivery model extends intervention support into home and telehealth settings and incorporates emotional, instrumental, informational, and appraisal support to influence self-efficacy, outcome expectancies, and social contexts relevant to tobacco and nicotine use during pregnancy.

#### 2.9.2 Overview of Intervention Arms

The *Smoke-Free HOPE* pilot feasibility trial tests two CHW-delivered intervention models: (a) CHW Brief Advice (CHW-BA) and (b) CHW Patient Navigation (CHW-PN). (Table 2). Both interventions consist of eight weekly (home/telehealth) sessions designed to provide proactive, personalized support for pregnant individuals using commercial tobacco products. Each arm offers equivalent CHW contact and is delivered alongside usual prenatal care, informed by best practices, including the 5 A’s and 5 R’s [1].

**Table 2.** Comparison of CHW-Brief Advice and CHW-Patient Navigation Intervention Components.

| **Component** | **CHW-Brief Advice (CHW-BA) Model** | **CHW-Patient Navigation (CHW-PN) Model** |
| --- | --- | --- |
| Behavioral Tobacco Cessation Approach | Structured “brief advice” based on the 5 A's [36] and the SCRIPT® (Smoking Cessation and Reduction in Pregnancy Treatment) program. CHWs present an educational video and introduce the self-help workbook “A Pregnant Woman’s Guide to Quit Smoking”[37] which promotes motivation and behavior change strategies. | Expanded patient navigation approach informed by the Ask-Advise-Connect framework [38]. CHWs guide participants through a CHW-curated list of state and community-based resources, including the *Kick-it CA* Quitline (www.kickitca.org), text- and app-based tools, and other evidence-based programs. Based on participant preferences, CHWs assist with selecting quit strategies, completing enrollment, and providing ongoing engagement, accountability, troubleshooting, and re-engagement support to promote continued use of selected cessation resources |
| Motivational Support | Use of motivational interviewing principles/techniques | (Same) |
| Home/Telehealth visits | Up to 8 home and telehealth visits | (Same) |
| Participant Incentives for participation only | Electronic gift cards | (Same) |
| Integration of CHW support typically reserved for pregnant patients with identified risk factors or SDOH needs[39] | CHW identifies unmet social needs through SDOH screening; assists with applications and linkage (e.g., food, housing, transportation resources as relevant); follows up weekly to confirm access, troubleshoot barriers, and update the care plan as needed. | (Same) |

Both CHW models are informed by SCT and the NIMHD Research Framework [34, 35]. The CHW-BA arm emphasizes skill-building through curriculum-based support, while the CHW-PN arm focuses on strengthening participants’ ability to access and use existing state- and community-based smoking cessation resources. Across both models, CHWs deliver directive and non-directive support that enhances behavioral capability and influences interpersonal and environmental factors relevant to quitting.

#### 2.9.3 CHW Workflow in Both Arms

Consistent with the American Public Health Association definition of CHWs as trusted frontline public health workers who bridge communities and social services, CHWs in the *Smoke-Free HOPE* trial are integrated into prenatal care to support tobacco cessation through home and telehealth visits. Both intervention models are grounded in evidence-based cessation strategies and adapted for CHW delivery. CHWs are classified as Level 2 according to Olaniran et al.’s typology [40] indicating paraprofessional workers with secondary education, formal standardized pre-service CHW certification, and salaried status within the health workforce.

CHWs operate within the LLUH care models and are oriented to a ‘whole person care’ approach. In addition to standardized CHW certification, they receive formal orientation through the LLUH Institute of Community Partnership on the Integrated CHW Care Model, social determinants of health, cultural competency, and trauma-informed care.

Integrated CHW services within LLUH follow federal guidelines for an 8-week treatment plan with weekly home or telehealth visits, each lasting about 60–90 minutes. Each visit includes EHR review, structured content delivery, participant check-ins, and documentation.

Consistent with current state-level guidelines and LLUH perinatal services, CHW referrals typically include pregnant patients and NICU admissions, with recognized behavioral risk factors or unmet health-related social needs on SDOH screening (e.g., housing instability or food insecurity) [39]. Tobacco use is recognized as an eligible risk factor; while not routinely used as a basis for referral at LLUH, the *Smoke-Free HOPE* trial is intentionally leveraging CHW referral pathways to address tobacco use during pregnancy.

The standardized perinatal CHW per-visit protocol is grounded in seven core functions: peer support; risk screening; self-management with problem-solving and goal setting; health education; referral and systems navigation; collaboration with the prenatal care team; and community outreach [41]. The integrated CHWs coordinate closely with the prenatal care team (e.g., social workers, public health nurses) to ensure continuity of care and timely referrals.

#### 2.9.4 Integration of Tobacco Cessation Module in Both Intervention Arms

These intervention strategies explicitly address pregnancy-specific cessation contexts, including household smoke-free rules, relapse risk across trimesters, and patterns of dual use involving e-cigarettes and combustible products, while leveraging CHWs’ strengths in social-needs navigation to support sustained engagement.

##### 2.9.4.1 CHW-Brief Advice (CHW-BA)

In this brief advice arm, the CHW will deliver the evidence-based *Smoking Cessation and Reduction in Pregnancy Treatment* (SCRIPT®) [37, 42] program using the 5 A’s framework for smoking cessation [36]. At the first home visit, the CHW introduces SCRIPT® by showing a video that builds self-efficacy, underscores maternal/fetal/infant risks, and models cessation strategies. Participants then complete the SCRIPT® workbook (*A Pregnant Woman’s Guide to Quit Smoking*; 5th–6th grade reading level; translated to Spanish), which covers cutting down, identifying reasons for quitting, understanding smoking cues, creating a stop-smoking contract, trigger identification, engaging a support “buddy,” stress-management (e.g., breathing exercises), managing urges/physical reactions, and coping/aversion techniques [37]. The materials were adapted for non-combustible tobacco products by adding discussion prompts and integrating supplemental handouts. The CHW serves as a health coach, scheduling and reinforcing workbook themes across the remaining seven weeks of the intervention and provides structured relapse support using SCRIPT® materials as needed. [See Multimedia Appendix 2 for an overview of weekly activities].

##### 2.9.4.2 CHW-Patient Navigation (CHW-PN)

In this arm, CHWs provide structured patient navigation support for tobacco cessation resources. CHWs introduce participants to a CHW-curated list of state and community-based cessation programs, including the Kick-it CA Quitline (www.kickitca.org), text- and app-based tools, and other local cessation supports. Based on participant preferences, the CHW helps participants select a quit strategy, complete enrollment in quit line program(s), and stay engaged through ongoing follow-up, accountability check-ins, and troubleshooting. Participants receive printed and digital materials, including the Kick-it CA “Quit Smoking Kit,” and support with downloading the Kick-it Smoking or Vaping application. CHWs monitor engagement throughout the program, address barriers, and re-engage participants as needed [See Multimedia Appendix 2 for an overview of weekly activities].

### CHW Training and Supervision

The *Smoke-Free HOPE* training program will prepare CHWs, all of whom will have completed a six-month certification program and the LLUH Institute Community Partnerships orientation program, to deliver both study interventions: Brief Advice (CHW-BA) and Patient Navigation (CHW-PN). Training will be conducted over three days, with Days 1–2 covering shared foundational content and Day 3 focused on arm-specific instruction.

#### 2.9.4.3 Days 1-2: Foundational Training (Both Arms)

The first two days will establish core knowledge in tobacco cessation and client-centered communication. On Day 1, CHWs will complete the Texas A&M *Tobacco Cessation for CHWs* course (4 CEUs)[43] and review the *National Community Health Worker Training Center CHW Toolkit: Tobacco Cessation[44]*. Trainers will contextualize the content using California and county-level maternal smoking data, review local cessation resources, and address evidence-based strategies for smoking cessation in pregnancy, including risks associated with secondhand smoke exposure. On Day 2, training will focus on reviewing and applying motivational interviewing (MI) and the Stages of Change framework. Through guided discussion and role-play based on maternal cessation scenarios, CHWs will strengthen MI skills and client engagement techniques relevant to both intervention arms.

#### 2.9.4.4 Day 3: Arm-Specific Instruction

On the final day, CHWs will receive intervention-specific training.

##### 2.9.4.4.1 Training: CHW-Brief Advice (CHW-BA)

CHWs will complete the SOPHE *SCRIPT® Training Bundle* (Modules 1–4[45], which covers screening for tobacco use and delivery of *SCRIPT®* counseling from initial assessment through follow-up. The session will include practice of *SCRIPT®* interactions, documentation, and integration of brief cessation counseling into prenatal and community care workflows. CHWs will receive the *SCRIPT® Guidelines and Counseling Manual for Prenatal Care* [42] for continued reference.

##### 2.9.4.4.2 Training: CHW-Patient Navigation (CHW-PN)

CHWs will be trained to link pregnant clients with local, state, and national cessation resources. Trainers will introduce *Kick It California* (https://kickitca.org/) and demonstrate how to navigate its services (e.g., quit-coach calls, text messaging support, self-help materials, mobile application). Role-play will reinforce motivational dialogue, navigation, and engagement strategies. Training will also cover documentation and follow-up procedures, and CHWs will be encouraged to contact local cessation programs and, when feasible, observe them to strengthen familiarity with available resources.

Across the three-day training, the *Smoke-Free HOPE* program is designed to equip CHWs with the competencies required to deliver both study interventions. Upon completion, CHWs will be able to: (a) deliver brief, evidence-based cessation counseling within the CHW–Brief Advice (CHW-BA) framework, and (b) effectively navigate and reinforce engagement with cessation resources within the CHW–Patient Navigation (CHW-PN) framework. In addition, CHWs will demonstrate culturally responsive communication, accurate referral practices, and capacity for sustained follow-up consistent with the *Smoke-Free HOPE* intervention protocol.

#### 2.9.4.5 Post-training Supervision Process Evaluation Procedures

Following CHW training, the PI provides structured supervision and debriefing during the early implementation phase to ensure consistency with the *Smoke-Free HOPE* protocol and to inform ongoing adaptation. Initially, the PI meets with each CHW following home or telehealth visits for the first 10 participants in each intervention arm to review visit content, confirm adherence to required components (e.g., tobacco use screening, counseling framework, documentation), and identify barriers to delivery or participant engagement. Feedback from these sessions is used to refine and tailor the intervention to participants’ cultural and pregnancy-related contexts. All supervisory interactions, field notes, and observations are documented in secure study files. The PI and program coordinator review these data during weekly team meetings to identify common challenges, support CHW learning, and document any adaptations made during implementation. The study leadership team discusses major protocol deviations or unanticipated adaptations and, if necessary, reports them to the IRB.

After implementing and supervising CHWs, the study will evaluate outcomes using a combination of quantitative and qualitative assessments, as described below.

### 2.10 Outcome and Process Evaluation

#### 2.10.1 Primary Feasibility and Acceptability Outcomes

The *Smoke-Free HOPE* pilot trial uses a mixed-methods evaluation process. Table 3 summarizes the primary outcomes, measures, and data sources.

**Table 3.**
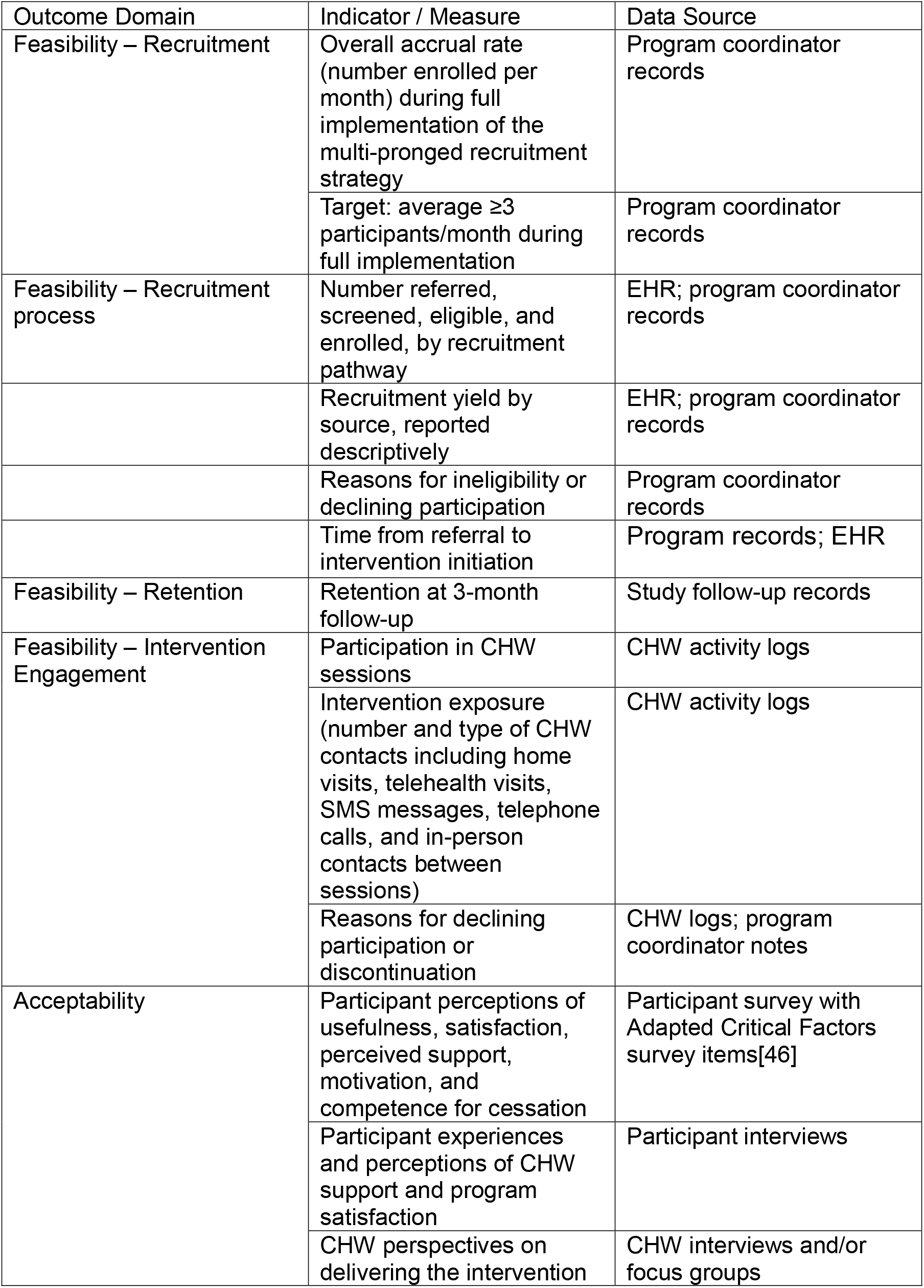
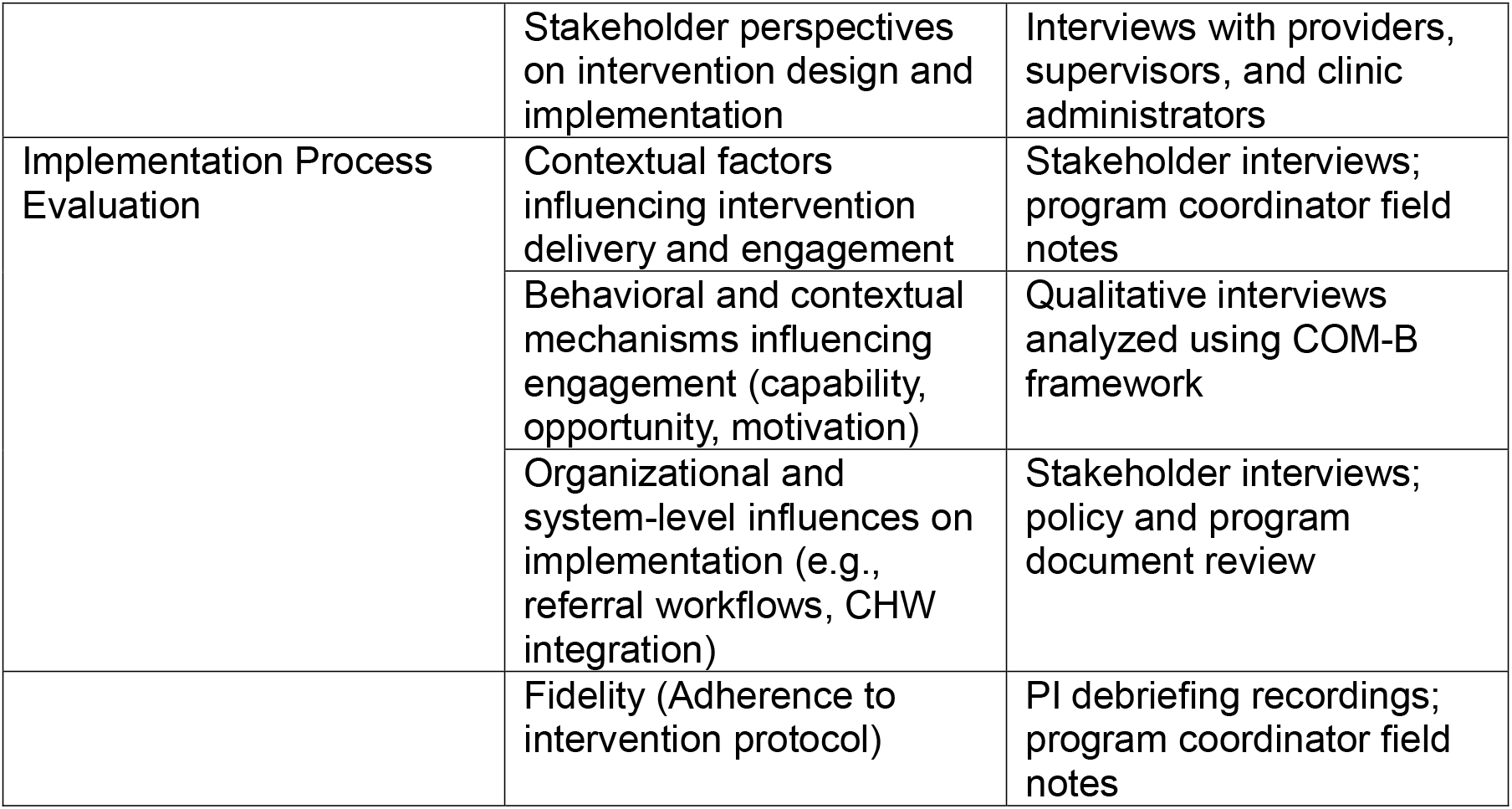
Implementation outcomes, measures, and data sources for the Smoke-Free HOPE pilot trial.

| Outcome Domain | Indicator / Measure | Data Source |
| --- | --- | --- |
| Feasibility – Recruitment | Overall accrual rate (number enrolled per month) during full implementation of the multi-pronged recruitment strategy | Program coordinator records |
| | Target: average $\geq 3$ participants/month during full implementation | Program coordinator records |
| Feasibility – Recruitment process | Number referred, screened, eligible, and enrolled, by recruitment pathway | EHR; program coordinator records |
|  | Recruitment yield by source, reported descriptively | EHR; program coordinator records |
|  | Reasons for ineligibility or declining participation | Program coordinator records |
|  | Time from referral to intervention initiation | Program records; EHR |
| Feasibility – Retention | Retention at 3-month follow-up | Study follow-up records |
| Feasibility – Intervention Engagement | Participation in CHW sessions | CHW activity logs |
|  | Intervention exposure (number and type of CHW contacts including home visits, telehealth visits, SMS messages, telephone calls, and in-person contacts between sessions) | CHW activity logs |
|  | Reasons for declining participation or discontinuation | CHW logs; program coordinator notes |
| Acceptability | Participant perceptions of usefulness, satisfaction, perceived support, motivation, and competence for cessation | Participant survey with Adapted Critical Factors survey items[46] |
|  | Participant experiences and perceptions of CHW support and program satisfaction | Participant interviews |
|  | CHW perspectives on delivering the intervention | CHW interviews and/or focus groups |
|  | Stakeholder perspectives on intervention design and implementation | Interviews with providers, supervisors, and clinic administrators |
| Implementation Process Evaluation | Contextual factors influencing intervention delivery and engagement | Stakeholder interviews; program coordinator field notes |
|  | Behavioral and contextual mechanisms influencing engagement (capability, opportunity, motivation) | Qualitative interviews analyzed using COM-B framework |
|  | Organizational and system-level influences on implementation (e.g., referral workflows, CHW integration) | Stakeholder interviews; policy and program document review |
|  | Fidelity (Adherence to intervention protocol) | PI debriefing recordings; program coordinator field notes |

##### 2.10.1.1 Feasibility

Feasibility will be evaluated across the domains listed in Table 3.

##### 2.10.1.2 Feasibility Criteria (Recruitment and Retention)

Progression to a future definitive trial will be guided by pre-specified recruitment and retention criteria, informed by methodological guidance recommending time-based recruitment targets and traffic-light progression rules for pilot and feasibility trials [19, 47, 48], and further informed by previous studies indicating challenges in recruiting prenatal populations with poverty-related risk factors [49]. The study-specific recruitment target of ≥3 participants/month was established based on anticipated accrual capacity after full implementation of all planned recruitment pathways. Recruitment feasibility will be based on this steady-state accrual rate, with ≥3 participants/month indicating feasibility (green/go), 2.0–2.9/month partial feasibility requiring refinement (amber/amend), and <2/month substantial feasibility concerns (red/reconsider). We will report recruitment during the staggered roll-out period and source-specific yields descriptively. Our retention rate criteria are informed by our previous work with the CTTP cohort of pregnant females, which indicated a 66% dropout rate for an on-site program (i.e., no telehealth component) [11]. Retention will be evaluated separately using the pre-specified criterion of >66%.

##### 2.10.1.3 Participant Engagement

Participant engagement with the intervention will be assessed based on participation in CHW-delivered sessions. Engagement will be measured by the number of CHW sessions participants complete within each intervention arm (60). ≥70% of enrolled participants completing five of the eight scheduled weekly CHW sessions will be considered indicative of acceptable intervention engagement [50]. CHWs and the program coordinator will document reasons for declining participation or discontinuing the program.

##### 2.10.1.4 Acceptability

Acceptability will be assessed using participant surveys and qualitative data. Participants will complete post-intervention surveys after CHW visit #8 and at program completion. Surveys will assess perceived impact of CHW interactions on motivation to reduce or quit tobacco or vaping, perceived competence related to cessation efforts, and satisfaction with CHW visits and communication.

Acceptability will be measured using an adapted version of the Critical Factors Survey [46], which assesses participant perceptions of program characteristics that support engagement and positive behavior change. Participants rate the importance of key program factors and how well the program achieved them using five-point Likert scales. The original survey included nine factors; for this study, the instrument was adapted to a 12th-grade reading level, and the item related to facilitating connection with culture was removed to align with the intervention context. The adapted version, therefore, includes eight items addressing strengths-based practice, relationship building, reliability and trust, non-judgmental support, boundaries and accountability, openness and honesty, opportunities for choice and self-motivation, and recognition of positive achievements.

Acceptability will also be explored qualitatively through interviews and focus groups with participants, CHWs, and other stakeholders to better understand perceptions of the intervention, satisfaction with CHW support, and recommendations for improvement.

##### 2.10.1.5 Secondary Exploratory Aims and Outcomes

The secondary aim of this pilot is to explore preliminary intervention efficacy on tobacco and nicotine use behaviors and related psychosocial outcomes among pregnant and postpartum participants. These measures will include: (a) Self-reported 7-day point prevalence abstinence; (b) Biochemical verification of abstinence (urine cotinine); (c) Reduction in tobacco use; (d) Quit attempts; (e) Engagement with cessation resources; (f) Perceived social support for quitting; and (g) Postpartum tobacco/nicotine use status.

#### 2.10.2 Process Evaluation

The process evaluation will examine how the *Smoke-Free HOPE* intervention is implemented in clinical and community settings and identify behavioral and contextual mechanisms that influence delivery, participant engagement, and perceptions of intervention acceptability. The process evaluation is guided by the COM-B, which serves as an analytic framework to examine behavioral and contextual mechanisms influencing both participant engagement with cessation support and CHWs’ delivery of the intervention [51–57].

Using this framework, qualitative data collection will primarily explore domains of reflective and automatic motivation (e.g., perceived usefulness, appropriateness, satisfaction, and emotional responses to the intervention) and social and physical opportunity (e.g., social support, clinic environment, workflow integration, and competing demands) that may influence engagement and satisfaction. We will also examine capability-related factors, including the clarity of intervention materials and the perceived adequacy of CHW training and skills.

Findings will be triangulated across data sources and integrated with quantitative feasibility results [51, 58, 59]. Fidelity will be assessed descriptively using CHW debriefs and program records to characterize adherence and variation in delivery.

##### 2.10.2.1 Qualitative Data Collection

Semi-structured interviews will be conducted with participants and CHWs to explore experiences with the *Smoke-Free HOPE* intervention. All participants will be invited to participate using a total population sampling approach to capture the full range of perspectives on intervention delivery and engagement. Interview guides will explore intervention delivery and receipt, perceived benefits and challenges of CHW-delivered cessation support, contextual influences on implementation, and recommendations for improvement.

The study will conduct informal key informant interviews with stakeholder groups, including CHW supervisors, referring healthcare providers (e.g., social workers and public health nurses), clinic administrators, and program leadership. Discussions will be documented in analytic memos that summarize implementation context and opportunities for refinement. To minimize potential social desirability bias, research staff independent of the intervention team will conduct interviews. Participants will be informed that responses are confidential and will not affect care or study participation. Interviewers will be trained in qualitative methods and will use a semi-structured guide to ensure consistency while allowing elaboration.

### 2.11 Sample Size and Rationale

This two-arm pilot feasibility trial is not powered to detect statistically significant differences between groups. Consistent with guidance from the NIDA Stage Model for early behavioral treatment development [60], which recommends approximately 15 participants per arm to evaluate feasibility parameters, the target sample size is 30 participants total (15 per arm). This sample will allow estimation of recruitment and retention [60]. Allowing for attrition, up to 40 participants may be enrolled to ensure that approximately 30 complete all study procedures. No formal power calculation was conducted.

### 2.12 Measures and Data Management

Table 1 provides an overview of assessments. Survey data will be collected verbally or electronically via a secure REDCap database. CHWs will document hard copies and when appropriate, enter reports into the EHR.

Primary outcomes are feasibility and acceptability; efficacy outcomes are exploratory to inform a future trial. Tobacco cessation will be assessed using 7-day point-prevalence abstinence, verified by urine cotinine (self-reported abstinence only) and self-report [61]. Samples will be collected at the LLUH outpatient laboratory and analyzed using LC–MS/MS [62]. Additional measures include tobacco use (quantity and frequency), quit attempts, and nicotine dependence using brief validated instruments across product types [22–25].

We will assess tobacco use patterns (e.g., product type, flavored products, switching) and history (initiation, prior quit attempts, pharmacotherapy use). Additional substance use includes cannabis [25] and alcohol (AUDIT-C) [25]. Social Cognitive Theory constructs (e.g., self-efficacy, motivation, perceived risks/benefits, social support, readiness to quit) will be assessed using adapted, validated measures [28, 63–65].

Clinical and contextual variables will be obtained from the EHR and participant report, including medical and psychiatric history, prenatal and birth outcomes, household smoking environment, and sociodemographic and mental health indicators [27].

Exploratory analyses will provide preliminary estimates of variability and potential effects to inform outcome selection and future trial design.

#### 2.12.1 Data Management and Security

The *Smoke-Free HOPE* program will manage all collected data using secure, standardized procedures to protect participant confidentiality and ensure compliance with institutional and federal data protection policies. Trial data will be entered directly into REDCap, a secure, cloud-based database serving as both the clinical chart and survey platform. Data entry will occur on password-protected, encrypted study laptops or tablets assigned to CHWs. All electronic data will be stored on firewall-protected servers maintained by LLUH, with REDCap enforcing role-based access control, automatic timestamping, and user authentication.

Any physical documentation related to CHW visits, key informant interviews, or focus groups, as well as audio recordings, will be stored in locked cabinets within the Principal Investigator’s (PI) secured office at the Loma Linda University School of Nursing. Keys will be kept in a separate locked drawer. All protected health information will be managed using a coded participant identification number (PID) system to remove direct identifiers. A master linkage file connecting names to PIDs will be maintained separately on a password-protected computer accessible only to the PI and designated study personnel. Signed consent forms, HIPAA authorizations, and incentive receipts will display only the PID.

Audio recordings will be labeled with PIDs and transcribed by a HIPAA-compliant transcription service approved by LLU Information Security. Transcripts will be de-identified and labeled solely by PID before analysis. All biospecimens and electronic survey data will likewise be linked by PID only; no personal identifiers will appear on laboratory samples or exported datasets.

#### 2.12.2 Data Retention and Destruction

De-identified research data (REDCap datasets, analysis files, and study documentation) will be retained on LLUH secure servers for at least 7 years after study completion or last publication, whichever is later, consistent with institutional policy and federal guidance. Audio recordings will be deleted after transcription is verified and quality checks are complete; only de-identified transcripts labeled by PID will be retained. Paper records (e.g., consent forms) will be stored in locked cabinets and retained for the same period, then destroyed by confidential shredding. Biospecimens will be retained only until planned assays and quality control are complete, then disposed of per LLUH biosafety procedures; no specimens will be used for secondary analyses without additional IRB approval and consent, if required. System backups will follow LLUH IT policies; access logs and role-based permissions will be reviewed periodically. Any future data sharing will use de-identified datasets under a Data Use Agreement and IRB/Privacy oversight.

### 2.13 Analytic Approach

Analyses emphasize feasibility and acceptability, with exploratory assessment of intervention effects. Consistent with a sequential mixed-methods design, quantitative trial data will be analyzed first to assess recruitment, engagement, retention, and acceptability, followed by qualitative analyses of contextual and implementation factors influencing these outcomes [15, 17]. Quantitative findings, qualitative findings, and CAB input will then be integrated to interpret findings, refine the intervention and procedures, and inform a fully powered trial in the future.

#### 2.13.1 Quantitative Analyses

Analyses will correspond to the pre-specified feasibility domains described in Table 3. We will summarize recruitment as the number of participants enrolled per month, separately for the recruitment start-up period and the full-implementation period. The primary recruitment feasibility estimate will be the average monthly accrual during the full implementation period, compared descriptively with the pre-specified traffic-light progression criteria. We will also summarize numbers referred, screened, eligible, and enrolled by recruitment source. We will summarize retention and other participant-level feasibility outcomes as proportions with 95% confidence intervals where appropriate. We will assess acceptability using the adapted Critical Factors Survey [46] and summarize results using Likert scales, with descriptive comparisons to prior CHW-led interventions where available.

Exploratory outcomes analysis will include effect-size estimation with 95% CIs. We will compare cotinine-verified 7-day point prevalence abstinence (PPA) at 1, 3, and 6 months between arms using risk ratios (RRs); RR ≥ 1.4 will be considered a heuristic signal of potential efficacy. Continuous abstinence with urine cotinine (Russell criteria) and time-specific PPA will also be estimated.

Longitudinal abstinence trajectories will be examined using GLMMs adjusted for baseline tobacco use/dependence. Secondary behavioral outcomes (e.g., cigarettes/day, quit attempts, dependence) will be summarized descriptively and analyzed for within-participant change and trajectories (paired tests, GLMMs) to inform variance estimates for a future trial.

Analyses will follow an intent-to-treat approach where feasible. Missing data will be addressed using multiple imputation with chained equations under missing-at-random assumptions, with sensitivity analyses including “missing = smoking.” Analyses will be conducted in R (R Foundation for Statistical Computing, Vienna, Austria) and SAS 9.4 (SAS Institute, Cary, NC).

#### 2.13.2 Qualitative Analyses

Qualitative data (interviews, focus groups, field notes, stakeholder data, and relevant documents) will be analyzed using a framework analysis informed by COM-B and Behavior Change Wheel constructs [51, 58, 59, 66, 67]. Analyses will examine behavioral and contextual mechanisms influencing participant engagement and CHW delivery of the intervention.

Coding will begin inductively within each data source to identify emerging themes and patterns, then be mapped deductively to COM-B domains to examine how contextual conditions and implementation processes influence feasibility (e.g., recruitment and retention), acceptability (e.g., perceived relevance and burden), and perceived mechanisms of impact (e.g., how CHW interactions or household environments shape cessation behaviors). This analytic process will identify recurrent themes, contextual variations, and modifiable determinants for future iterations of the intervention.

Two researchers will code transcripts independently, resolving discrepancies through consensus. Data will be managed in NVivo (Lumivero, version 14). The audit trail will document coding decisions and theme development to support analytic transparency and rigor. Findings will be interpreted across data sources to better understand contextual influences on implementation, including factors related to maternal smoking cessation and CHW integration within primary care and community settings.

#### 2.13.3 Data Integration

Integration will occur at two stages: (a) during qualitative data collection, where preliminary quantitative findings inform refinement of interview guides, and (b) during interpretation through structured joint display tables that compare quantitative results with qualitative themes. This sequential synthesis will yield a theory-informed understanding of how contextual and implementation factors shape study feasibility, participant engagement, and potential mechanisms of impact.

CAB input from the development and implementation phases will be used to contextualize interpretation of primary outcomes, assess the relevance of barriers and facilitators, and prioritize feasible refinements to intervention components, CHW training, and implementation procedures. This approach will generate actionable recommendations to refine the intervention, optimize implementation, and inform future scale-up.

### 2.14 Ethics, Oversight, and Monitoring

#### 2.14.1 Ethical Considerations and IRB Approval

The Loma Linda University Institutional Review Board (IRB) approved the *Smoke-Free HOPE* study protocol as minimal risk (Approval No. 5220112). All research activities comply with federal regulations (45 CFR 46), institutional policies, and the Belmont Report.

All study materials (e.g., recruitment, consent, training, and survey materials) and participant compensation were approved by the IRB. We will submit protocol modifications to the IRB for approval before implementation.

All research staff, including CHWs, completed IRB and HIPAA training. Participants will be informed that participation is voluntary and will not affect their medical care or access to community services.

#### 2.14.2 Adverse Event Reporting and Participant Safety

Adverse events (AEs) and serious adverse events (SAEs) will be identified, documented, and reported throughout participation using a standardized process. Study staff will assess AEs at each visit (home or telehealth) and during follow-up contacts; participants may also report events via a dedicated study contact. AEs include any adverse medical, psychological, or social event; SAEs include events resulting in hospitalization, life-threatening conditions, disability, congenital anomaly, or death.

The PI and Medical Director will review all events for severity, expectedness, and relatedness. Anticipated events are primarily minor (e.g., transient mood changes, nicotine withdrawal symptoms, headache, dizziness, or emotional distress during counseling). All AEs and SAEs will be recorded using standardized forms that capture onset, duration, severity, outcome, and actions taken.

SAEs related to the intervention will be reported to the IRB within 3 business days; other SAEs within 15 days. Participants experiencing serious or unexpected intervention-related events will discontinue the intervention phase for safety monitoring but will remain in follow-up when feasible. Events will be followed to resolution or stabilization, with referral as needed. The Data and Safety Monitoring Committee (DSMC) will review summaries and report them annually to the IRB.

#### 2.14.3 Data and Safety Monitoring Committee and Trial Oversight

Monitoring includes ongoing safety reporting and oversight by a Data and Safety Monitoring Committee (DSMC). The trial is registered at ClinicalTrials.gov (NCT06438549). Reporting follows the SPIRIT Statement and the CONSORT extension for pilot and feasibility trials, applied as appropriate to an early-phase pilot [19–21]. Table 1 summarizes enrolment, intervention, and assessment schedules for the pilot RCT and process evaluation.

## 3 Dissemination and Knowledge Translation

The CAB will support dissemination by co-developing plain-language summaries, community briefings, and CHW/clinic toolkits. CAB members may co-present findings to local health systems and community forums and advise on policy-relevant messaging (e.g., CHW reimbursement and integration) to ensure outputs are accessible, actionable, and aligned with community priorities. Findings will also be presented at conferences and published in peer-reviewed journals.

## 4 Results

Between 2022 and 2024, the CAB met multiple times and provided feedback that informed the intervention, recruitment strategies, study procedures, and feasibility and acceptability assessments.

In San Bernardino County, recruitment occurred over approximately 18 months, from 2024 to 2026, and concluded in May 2026. Follow-up activities are nearing completion, while data management and preliminary analyses are underway. Study findings are expected to be submitted for publication in 2026.

Several protocol modifications were made during implementation, informed by CAB input, pretesting, and implementation experience. Key amendments and their rationale are summarized in Multimedia Appendix 3.

## 5 Discussion

### 5.1 Summary of Contribution

Building on findings from our prior maternal tobacco cessation program evaluation, the *Smoke-Free HOPE* trial incorporates CHWs to address persistent challenges in engagement, retention, and relapse observed in earlier models. Consistent with the American Public Health Association definition of CHWs as trusted frontline public health workers who serve as a bridge between communities and social services, CHW-based tobacco cessation interventions using tailored messaging have shown reach among general and disparity populations in the U.S. [68–76], including adults in rural underserved regions [76, 77], African American/Black women [78], adults with serious mental illness [75], and those with chronic conditions [73]. However, much of the evidence on formally trained CHWs delivering cessation interventions, particularly in perinatal populations, comes from studies conducted outside the U.S. (e.g., Hong Kong, India, Thailand, Vietnam) (24-29).

The *Smoke-Free HOPE* trial protocol is designed to extend this literature and, to our knowledge, is the first to integrate a CHW-delivered cessation intervention tailored to pregnancy in the US. The intervention adapts the SCRIPT® curriculum for CHW delivery across home and telehealth visits. It is designed for the contemporary tobacco and nicotine landscape in pregnancy, accounting for combustible products and non-combustible products and patterns of dual use without privileging a single cessation pathway [37, 42]. Importantly, the intervention is tailored to pregnancy-specific contexts, including variability in cessation goals, to account for scenarios where some patients may use non-combustible tobacco products for cessation, some may be dual users, and others may want to achieve complete nicotine cessation [79–81]. By focusing on pregnancy-specific contextual factors and leveraging CHW-supported navigation to community and health system resources, this approach builds on and extends prior CHW cessation models to a perinatal context.

### 5.2 Anticipated Challenges and Mitigation Strategies

Due to well-documented under-reporting of tobacco use and tobacco-related stigma in pregnancy [82–84], and additional challenges associated with pregnancy, recruitment, and retention of pregnant individuals who use tobacco products are anticipated to be the most significant challenges. However, this study builds on lessons learned from prior maternal smoking-cessation services provided by LLUH (CTTP) and on the established referral base. To enhance reach and engagement, the study uses multiple recruitment pathways, including electronic health record screening and invitations, provider referrals, and community outreach; CHWs will implement the intervention using tailored materials and flexible scheduling (telehealth or home visits). Additionally, small, activity-linked incentives and brief check-ins by CHWs and the program coordinator are intended to support adherence without undue influence.

The study design requires separate CHWs for each intervention arm to minimize contamination and preserve intervention integrity, which may introduce hiring and staffing challenges. To address this, the LLUH ICP will leverage existing infrastructure and experience managing grant-funded programs to support targeted recruitment, role delineation, and workload allocation. Variability in CHW delivery represents an additional risk and will be mitigated through standardized training, ongoing supervision, and structured monitoring of intervention delivery using checklists, electronic documentation, and regular debriefs. These procedures are intended to support consistency while allowing appropriate contextual adaptation within a pilot feasibility framework.

Participant burden may affect data completeness, particularly for biospecimens used to verify abstinence. To mitigate this risk, the program coordinator will work closely with participants to schedule surveys at convenient times and provide flexible specimen collection options. Integration within busy prenatal workflows may also pose logistical difficulties; EHR templates, multiple referral options (e.g., a referral menu and a website), and tip sheets for referring providers will mitigate these challenges.

Finally, social and structural barriers such as housing instability, food insecurity, and co-occurring substance use may compete with cessation goals. The evolving tobacco and nicotine landscape may complicate cessation trajectories, participant goals, and outcome measurement; this is addressed through flexible, participant-centered cessation planning and adapted assessment tools that accommodate product switching and dual use. The intervention addresses these challenges through CHW-led screening and navigation to community resources, as well as trauma-informed, strengths-based support.

### 5.3 Implications and Future Research

*Smoke-Free HOPE* integrates behavioral and social support within CHW-led perinatal care to improve engagement, retention, and long-term cessation among populations at highest risk for adverse birth outcomes. Findings from this pilot feasibility trial will inform the refinement of intervention components, implementation strategies, and outcome selection for scale-up within FQHC networks, as well as the design of a future RCT to advance maternal and infant health equity through sustainable, community-driven cessation approaches. This study will also assess the feasibility of integrating CHW-led tobacco cessation into existing CHW/perinatal support services for high-risk pregnancies [9, 85, 86].

## List of Abbreviations

| Abbreviation | Definition |
| --- | --- |
| 5 A's and 5 R's | 5A (Ask, Advice, Assess, Assist, Arrange) and 5R (Relevance, Risk, Reward, Roadblock, Repetition) |
| AEs | Adverse events |
| AUDIT-C | Alcohol Use Disorders Identification Test-Consumption |
| CAB | Community Advisory Board |
| CHW | Community Health Worker |
| CHW-BA | CHW-Brief Advice Intervention Arm |
| CHW-PN | CHW-Patient Navigation Intervention Arm |
| COM-B | Capability, Opportunity, Motivation-Behavior framework |
| CONSORT | Consolidated Standards of Reporting Trials |
| CTTP | Comprehensive Tobacco Treatment Program |
| DSMC | Data and Safety Monitoring Committee |
| EHR | Electronic Health Record |
| FQHC | Federally Qualified Health Centers |
| FTCD | Fagerström Test for Cigarette Dependence |
| GLMMs | Generalized Linear Mixed-Effects Models |
| HIPAA | Health Insurance Portability Act |
| LLUH | Loma Linda University Health |
| m-CEQ | Modified Cigarette Evaluation Questionnaire |
| NIDA | National Institute on Drug Abuse |
| NIMHD | National Institute on Minority Health and Health Disparities |
| NRT | Nicotine Replacement Therapy |
| PC | Program Coordinator |
| PID | Participant Identification Number |
| PPA | Point Prevalence Abstinence |
| RCT | Randomized Controlled Trial |
| REDCap | Research Electronic Data Capture web-based application |
| RIB | Institutional Review Board |
| RR | Risk Ratios |
| SAEs | Serious Adverse Events |
| SCT | Social Cognitive Theory |
| SCRIPT | Smoking Cessation and Reduction in Pregnancy Treatment |
| SDOH | Social Determinants of Health |
| SPIRIT | Standard Protocol Items: Recommendations for Interventional Trials |

## Declarations

### Ethics approval and consent to participate

Ethical approval has been granted by the Loma Linda University Health Institutional Review Board (Approval No. 5220112), was in place prior to commencing the study.

### Consent for publication

Non-applicable

### Availability of data and materials

The data supporting this research are available from the authors on reasonable request.

### Author contributions

ABP and PNS drafted the initial protocol. BO designed the maternal tobacco cessation program (CTTP) that informed the intervention. JT provided subject matter expertise in intervention development and community health worker (CHW) training. JCB developed the LLUH integrated CHW model and guided integration of the CHW role within the study. BR supported intervention refinement and coordinated data collection. BO and RY serve as medical directors and contributed to recruitment strategy development. All authors contributed to the conception, design, and revision of the final protocol.

## Supporting information

Multimedia Supplemental File 1

Multimedia Appendix 2

Multimedia Appendix 3

## Data Availability

All data produced in the present study are available upon reasonable request to the authors.

## Acknowledgements

We thank the *Smoke-Free HOPE* Community Advisory Board members for their sustained guidance and partnership in developing and implementing this study. We are grateful to the leadership and clinical team at Loma Linda University Health Maternal– Fetal Medicine for ongoing support in integrating study procedures into clinical operations. We acknowledge the LLUH Institute for Community Partners Community Health Worker (CHW) Program for collaboration on CHW training, supervision, and implementation, and the *Smoke-Free HOPE* Program Coordination Team for project management, recruitment, and operational support.

