## Supplementary material for "Feasibility of a Community Health Worker-delivered Tobacco Cessation Intervention During Pregnancy: Protocol for the Smoke-Free HOPE Pilot Randomized Controlled Trial": Multimedia Appendix 2

**Overview of CHW Activities Across Eight Weekly Visits in both arms: Arm A (CHW–Brief Advice) vs. Intervention Arm B (CHW–Patient Navigation)**

**Notes:** Both arms include rapport building, ongoing encouragement, monitoring quit status, and relapse support. Arm A is curriculum‑driven, using SCRIPT® video/workbook and behavioral exercises; Arm B adapts the Ask–Advise–Connect [1] approach to link participants to a CHW-curated set of state, community, and digital cessation resources and to provide navigation and maintenance support.

| **Visit** | **Arm A: CHW Brief**  **Intervention (CHW-BA)** | **Arm B: CHW Patient Navigation (CHW-PN)** |
| --- | --- | --- |
| 1  Home Visit  (~60 minutes) | Build rapport; assess tobacco/nicotine use; introduce the structured SCRIPT® curriculum (video + workbook)[2, 3]; discuss health effects and motivations; support quit-date planning and complete quit agreement; encourage identification of social support; introduce smoking diary; offer referrals for household smokers. | Build rapport; assess tobacco/nicotine use; introduce curated cessation resources using the Ask–Advise–Connect approach; provide Kick-It CA materials/app; help select and enroll in preferred resources; support quit-date planning; offer referrals for household smokers. |
| 2 (Pre-quit date)  Home or Telehealth Visit  (~30 minutes) | Review the smoking diary; reinforce reduction or scheduled smoking strategies; introduce deep-breathing and aversion techniques; help identify a 'stop-smoking buddy'; prepare a SCRIPT®-based quit plan, including environmental changes and coping tools; offer encouragement and early troubleshooting. | Follow up on engagement with selected cessation resources; assist with enrollment if not completed; troubleshoot early barriers; reinforce quit-date plan; provide encouragement. |
| 3 (Quit date)  Telehealth Visit  (~15-20 minutes) | Assess progress; reinforce SCRIPT® behavioral strategies; review coping skills (4Ds); celebrate progress; support buddy system; provide early problem-solving for cravings or withdrawal. | Assess progress; reinforce benefits of quitting; confirm engagement with Quitline, community, and/or digital supports; provide navigation and problem-solving support; celebrate progress; re-engage participant with selected resources as needed. |
| 4 (Post‑quit)  Home or Telehealth Visit  (~15-20 minutes) | Assess quit status or slips; address withdrawal symptoms; troubleshoot challenges; reinforce SCRIPT® coping tools; re-engage participant with behavioral strategies as needed; celebrate ongoing efforts. | Assess quit status; if relapsed, support selecting a new quit date; troubleshoot challenges; re-engage participant with preferred cessation resources; celebrate ongoing efforts; reinforce continued engagement. |
| 5 (Post‑quit)  Home or Telehealth Visit  (~15-20 minutes) | Reassess progress; apply SCRIPT® relapse-prevention strategies (triggers, social situations, self-care, buddy support); troubleshoot any challenges; strengthen self-efficacy; provide behavioral maintenance support; affirm progress. | Monitor engagement with cessation resources; troubleshoot barriers; offer navigation-based maintenance support; re-engage or adjust selected quit pathway as needed; strengthen self-efficacy; affirm progress. |
| 6 (Post‑quit)  Home or Telehealth Visit  (~15-20 minutes) | Reassess progress; reinforce coping strategies and self-efficacy; address new or ongoing triggers; continue strengthening alternative behaviors; affirm ongoing commitment. | Assess continued engagement with Quitline, community programs, and digital tools; troubleshoot challenges; provide accountability and re-engagement support; affirm ongoing commitment. |
| 7 (Post‑quit)  Home or Telehealth Visit  (~15-20 minutes) | Assess slips or continued abstinence; review progress; celebrate achievements; reinforce long-term SCRIPT® maintenance strategies; bolster confidence in remaining smoke-free. | Check quit status and engagement; review progress; celebrate achievements; support long-term maintenance planning using resources; bolster confidence; re-engage participant if support has lapsed. |
| 8 (Post‑quit)  Home or Telehealth Visit  (~15-20 minutes) | Review and celebrate the overall quit journey; summarize long-term SCRIPT® behavioral maintenance strategies; review community cessation supports; make referrals as needed; reinforce self-efficacy for sustained abstinence. | Review and celebrate the overall quit journey; ensure connection to long-term cessation supports (Quitline, community programs, digital tools); assist with final referrals; reinforce self-efficacy for sustained abstinence. |

1.	Vidrine, J.I., et al., *Ask-Advise-Connect: a new approach to smoking treatment delivery in health care settings.* JAMA Intern Med, 2013. **173**(6): p. 458-64.

2.	Windsor, R., *SCRIPT: Guidelines and Counseling Manual for Prenatal Care*. 2014. p. 1-16.

3.	Windsor, R.A., *A Pregnant Woman's Guide to Quit Smoking*. 1997, Birmingham, AL: Quality Press, INC.
