## Supplementary material for "Feasibility of a Community Health Worker-delivered Tobacco Cessation Intervention During Pregnancy: Protocol for the Smoke-Free HOPE Pilot Randomized Controlled Trial": Multimedia Appendix 3

**Summary of Key Protocol Amendments During Study Implementation**

| **Date** | **Major protocol amendment** | **Rationale and implications** |
| --- | --- | --- |
| **August 2024** | **Broadened participant eligibility.** Eligibility was expanded beyond cigarette smoking to include other tobacco and nicotine products, and the <24-week gestational-age restriction was removed. | Informed by CAB input during intervention development, eligibility was broadened to better reflect tobacco and nicotine use during pregnancy and expand recruitment reach. Subsequent amendments aligned recruitment language and procedures with the revised eligibility criteria. |
| **December 2024** | **Removed the usual-care arm and expanded recruitment beyond LLUH prenatal care.** Participants were no longer required to receive prenatal care at LLUH, allowing direct referrals from providers and individuals outside the LLUH system. | The study design and recruitment pathways were modified in response to implementation and recruitment considerations, allowing broader participation beyond the original LLUH prenatal-care setting. |
| **January–August 2025** | **Expanded recruitment** through community-based organizations and providers, introduced an EHR research referral order, and added social media recruitment. | These changes expanded referral and recruitment pathways and strengthened connections to community resources. An EHR system update also created an opportunity to add *Smoke-Free HOPE* to the electronic referral menu, providing an additional provider-based referral pathway. |
